# Effects of stricter management guidelines on return-to-play timeframes following concussion in professional Australian Rules football

**DOI:** 10.1101/2021.01.25.21250431

**Authors:** Alan J Pearce, Doug A King, Adam J White, Catherine M Suter

**Affiliations:** College of Science, Health and Engineering, La Trobe University, Melbourne, Australia; Sports Performance Research Institute New Zealand (SPRINZ), Faculty of Health and Environmental Science, Auckland University of Technology, Auckland, New Zealand; Department of Sport, Health Sciences and Social Work, Faculty of Health and Life Sciences, Oxford Brookes University, Oxford, Britain; Department of Neuropathology, Royal Prince Alfred Hospital, Sydney Australia

**Keywords:** Sports concussion, Injury expectations, Mild traumatic brain injury, Return to play

## Abstract

**Background:** Management of concussion remains a serious issue for professional sports, particularly with the growing knowledge on the consequences of repetitive concussion. One primary concern is the subjective assessment of recovery that dictates the time until a concussed athlete is returned to competition. In response to this concern, the Australian Football League (AFL) changed its policy in 2020 such that clearance for return-to-play was extended from one day, to a minimum of five days, prior to the next scheduled match.

**Objective:** We sought to examine the impact of the AFL policy change by asking whether the time to return-to-play after concussion was increased in the 2020 season relative to previous years.

**Methods:** Retrospective data on injury and return-to-play were sourced from publicly available tables published on the AFL website. We compared the number of matches missed and the number of days missed in concussed players across 2017 to 2020 inclusive.

**Results:** Analysis of data from 166 concussed players revealed no increase in the number of matches missed in 2020 relative to previous years as would have been expected from an extend recovery protocol. Considering the number of days missed in 2020 relative to 2017-19 we found, paradoxically, that there was an overall *reduction* in the average time to return-to-play in 2020 (11.2 *vs* 16.2 days).

**Conclusion:** This study demonstrates that any policy change around concussion management requires ongoing auditing to ensure clearance meets policy objectives and highlights the need for objective measures for return-to-play after concussion.

## 1.0 Introduction

The international consensus statement on the management of concussion in sport [1] promotes a strategy whereby an athlete recovering from concussion is exposed to graduated stages of increasing activity, only passing from one stage to the next if the concussion symptoms are resolving. A minimum rest time of 24 to 48 hours post-concussion is recommended prior to beginning the return-to-play steps, and the minimum recommended interval between stages is no less than 24 hours. Clearance for return-to-play is a medical judgement made by the team doctor in consultation with the coach and player. Clearance can only occur if the athlete declares that they are symptom free, and the treating physician deems the athlete is safe to return to sport [1].

This seems a sensible policy, however there are criticisms of this approach, the most prominent of which is the reliance on self-reporting of symptoms for progression through the various stages. This subjectivity in assessing recovery is further complicated by the fact that after a concussion, symptom resolution and an individual’s physiological functioning are not necessarily correlated [2]. Consequently, there are ongoing concerns that athletes are returning to competition prematurely after a concussion, increasing both the risk of further injury, and the likelihood of persistent post-concussion symptoms [1, 3-6].

In an attempt to address these concerns, the Australian Football League (AFL), who adhere to the protocols based on the Berlin recommendations, [7] recently changed their concussion management guidelines. Prior to the 2020 season, return-to-play clearance stipulated AFL players be cleared by the team doctor prior to the next match; in theory a player could be cleared the day before the next match. Now in 2020, the AFL have introduced a stricter policy, mandating medical clearance occur at least five days prior to the team’s next scheduled match [8, 9]. The AFL proposed this policy change would make it more difficult for players to return-to-play the week after sustaining a concussion, however these changes fall short of mandating a compulsory break in play for at least one competition match.

The effectiveness of this strategy to increase the time between concussion and return-to-play can be assessed objectively, because the AFL publishes detailed information on injuries and play via its website. We interrogated this publicly available data to test the hypothesis that the new concussion management policy would increase the time players were out of the game in the number of matches missed, and the overall number of days missed between concussion and return to competition.

## 2.0 Methods

Concussion injury and fixture data were collected from public internet websites www.afl.com.au/matches/injury-list and www.afl.com.au/fixture, respectively. Inclusion/exclusion criteria are shown in **Figure 1**. Players were included if they were: 1) AFL listed; 2) played at least one match in the AFL; 3) were included in the Injury List with concussion and; 4) either returned-to-play within the season or were indefinitely removed from play because of concussion. Ethical consent was sought from the Health and Disability Ethics Committee NZ but was not required. Using de-identified athlete data, players who returned-to-play after concussion had the number of matches missed recorded, and also the days between the concussion injury and the next match played. We surveyed this data for the years of 2017 to 2020 (inclusive). Due to the COVID-19 pandemic, the number of rounds in the 2020 competition was reduced from 23 rounds in 2017-19 seasons, to 18 rounds in 2020. Consequently, data from 2017-19 on athletes who returned-to-play following a concussion after round 18 were not included in the analysis.

**Figure 1.**
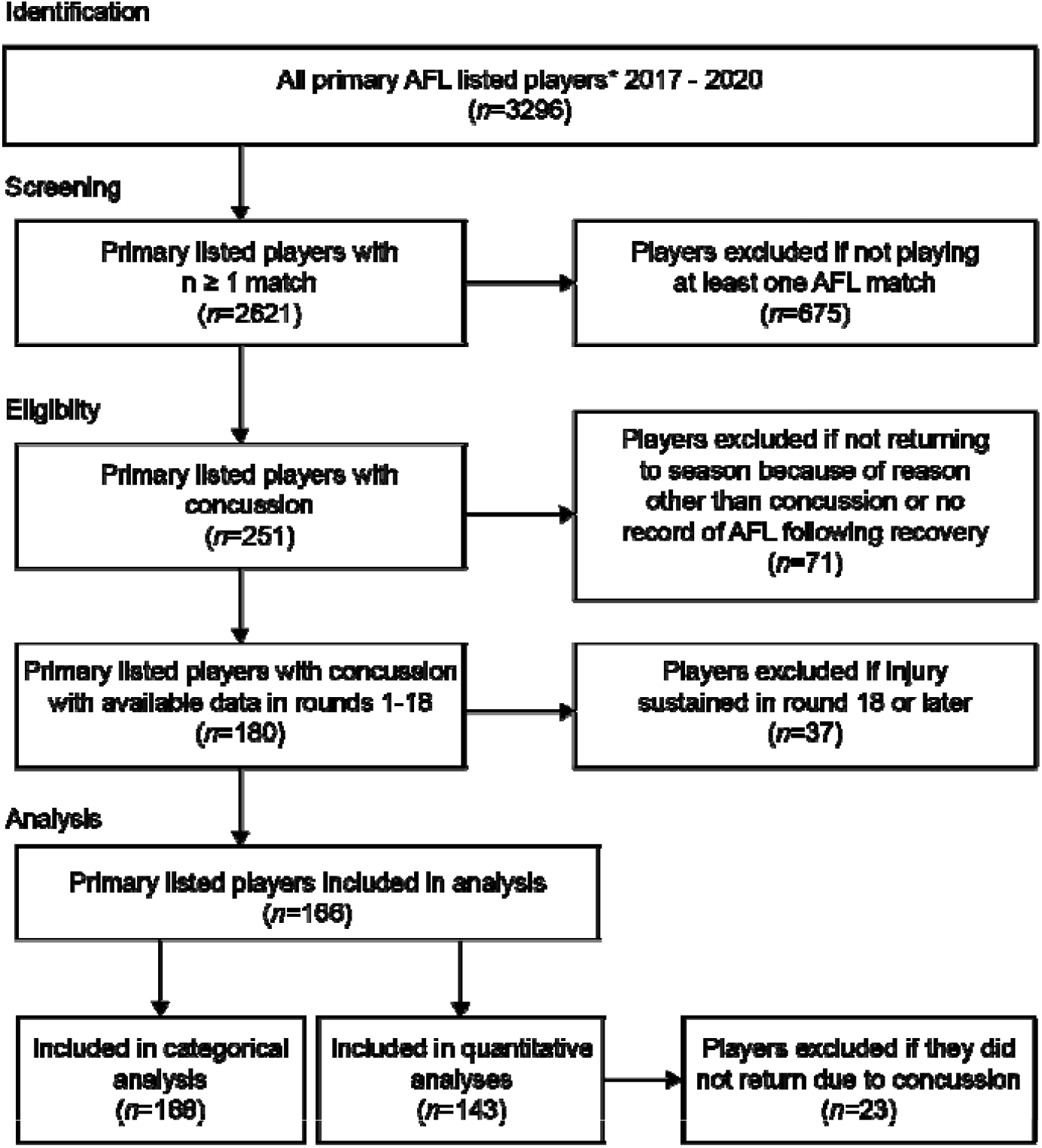
Data selection process for inclusion of athletes for analysis.

Following screening of data, statistical comparisons of quantitative data across all years were made using the nonparametric independent-samples Kruskal-Wallis test; the Mann-Whitney U Test was used to compare 2020 to all other years combined. Contingency tables were used to compare categorical data with Chi-Square tests for comparison between 2020 and all other years, and Fisher’s Exact test for 2020 versus all years combined. Data are presented as mean and 95% confidence intervals (CI), and where appropriate, effect size (Cohen’s *d*) was used to calculate effect differences between groups (≤0.2 = small; 0.31-0.8 = medium; ≥0.81 = large) [10]. Kaplan–Meier estimators were used to determine return-to-play probabilities and these were compared using the Mantel-Cox log rank test. For all tests, an alpha value of 0.05 was considered significant.

## 3.0 Results

As outlined above, the timeline for the 2020 season was disrupted by COVID-19 such that meaningful comparisons can only be made considering 18 rounds of play. We began comparisons by compiling the details of concussions sustained across the 18 rounds of 2020, along with first 18 rounds of each of the 2017 to 2019 seasons. The incidence of concussion was similar across the years (mean 42, range 36-48). **Table 1** shows the descriptive return-to-play statistics for each year.

**Table 1.** Descriptive statistics for concussion and return-to-play in professional AFL from 2017 to 2020

|  | **2017** | **2018** | **2019** | **2020** |
| --- | --- | --- | --- | --- |
| **Players concussed in rounds 1-18 *n*** | 43 | 48 | 39 | 36 |
| **Sum of matches missed<sup>a</sup> *n*** | 29 | 61 | 55 | 27 |
| **Mean matches missed<sup>a</sup> *n* (±SD)** | 0.8 (1.1) | 1.5 (2.4) | 1.7 (1.9) | 0.8 (1.0) |
| **Total time to RTP<sup>a</sup> *days*** | 453 | 710 | 618 | 370 |
| **Mean time to RTP<sup>a</sup> *days* (±SD)** | 12.9 (8.1) | 16.9 (17.2) | 18.7 (13.7) | 11.2 (5.3) |
| **Players not returning to season<sup>b</sup> *n* (%)** | 8 (18.6) | 6 (12.5) | 6 (15.0) | 3 (8.3) |
| **Players not missing a match after concussion *n* (%)** | 18 (41.9) | 18 (37.5) | 11 (28.2) | 16 (44.4) |
| **Players missing 10 or fewer days after concussion *n* (%)** | 18 (41.8) | 20 (41.7) | 11 (28.2) | 20 (55.6) |

<sup>a</sup> Only counted for those players returning to play in that season; <sup>b</sup> Not returning because of concussion; RTP, return-to-play; SD, standard deviation

In quantitative analysis of 143 players, we found no difference in the mean number of matches missed after a concussion between all years (Kruskal-Wallis, *p* = 0.101), and also no difference within the years of the previous return-to-play policy (2017/18/19; *p* = 0.176), (see **Figure 2a**). We thus considered 2017-19 (previous policy) as a group and found that there was reduction in the mean number of matches missed in 2020 relative to the previous years combined (0.8 vs 1.3), although this was not statistically significant, we found a moderate decreased effect size (Mann-Whitney U, *p* = 0.391; *d*=-0.345), (see **Figure 2b**). This finding was unexpected as the new policy was designed to *increase* the time to return-to-play after a concussion.

**Figure 2.**
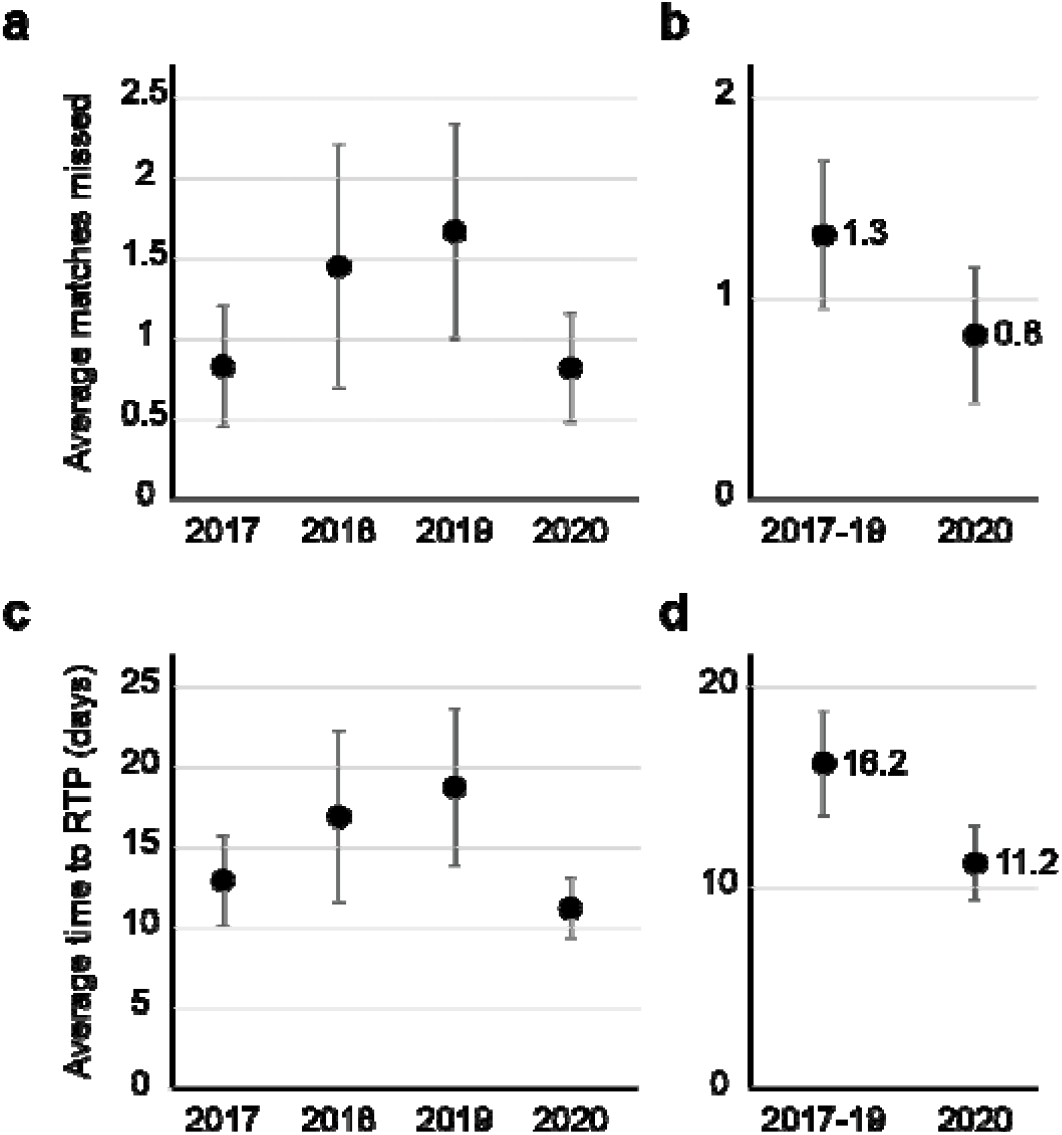
Matches missed after concussion (**a, b**) and time to return-to-play (**c, d**) across the years indicated. Only players who did return to the same season are included. Plotted is group mean ±95 CI. (RTP: return-to-play; 2017 *n* = 35, 2018 *n* = 42, 2019 *n* = 33, 2020 *n* = 33).

We considered that these results might be attributable to changes in match scheduling because of COVID-19; some matches in 2020 were delayed, or brought forward. We thus compared the *time* to return-to-play (in days) after concussion (see **Table 1**). We found that across all rounds that there were no significant differences observed in mean return-to-play times, although 2020 had the least number of days missed across all years, (see **Figure 2c**). There were no significant differences observed between years with the old policy, so we again considered 2017-2019 as a group, and compared the time to return-to-play to 2020. These results, although not statistically significant, show that there was a large reduction in time to return-to-play in 2020 compared to the previous years (11.2 vs 16.2 days; *d*=0.524).

There were 23 additional players with a concussion that were indefinitely sidelined because of their concussion injury across the four years (see **Table 1**). These players could not be included in the quantitative analysis above but could be included in a categorical analysis of return-to-play. We categorised all 166 players as either having missed no match after their injury, or one or more matches. While the proportion of players not missing a match after concussion was highest in 2020 (44%) there was no significant difference between years, or between 2020 and all years combined (see **Figure 3a**).

**Figure 3.**
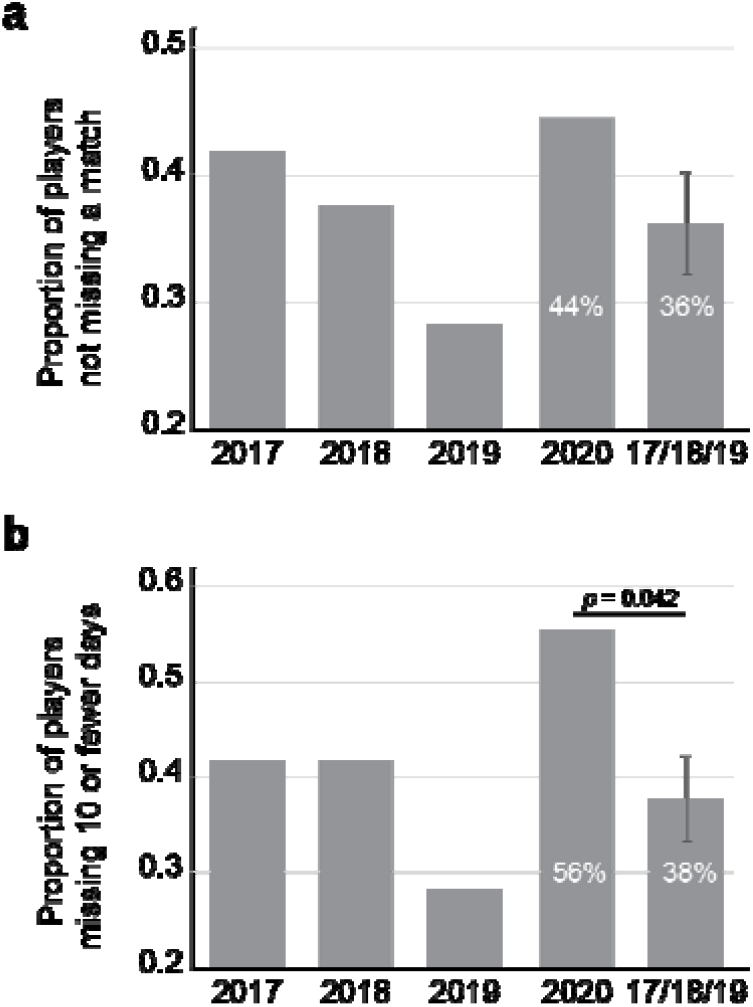
Histograms showing the proportion of all players categorised as (**a**) having not missed a match after concussion and (**b**) having missed 10 or fewer days of play after a concussion. Error bars in grouped data from 2017-2019 represent ±95 CI. (2017 *n* = 43, 2018 *n* = 48, 2019 *n* = 39, 2020 *n* = 36).

We also categorised the time to return-to-play by categorising players into those missing 10 or fewer days, and those missing more than 10 days after their concussion. In 2020, more than half of all players (56%) who sustained a concussion were back on the field within 10 days (see **Figure 3b**). Again, there was no statistical differences observed between all years, or years of the prior policy. However, the increase in the proportion of players taking 10 days or less to return in 2020 was significantly reduced in 2020 relative to 2017-19 (χ^2^(1, *n*=166) = 3.701, Fisher’s Exact *p* = 0.042; see **Figure 2d**).

The analyses above rely on comparing the distribution of means using nonparametric tests. A more holistic view of the data can be taken by comparing probabilities of returning to play. We thus generated Kaplan Meier (KM) plots to show the relative rates of return-to-play after concussion across the years. **Figure 4a** shows KM plots for each year individually, with 2020 showing the fastest trajectory for all players returned. When grouping the years of previous policy together, it is clear that 2020 had a significantly faster trajectory to return-to-play than the years of the previous policy combined (Mantel-Cox log rank test, *p* = 0.040; see **Figure 4b**).

**Figure 4.**
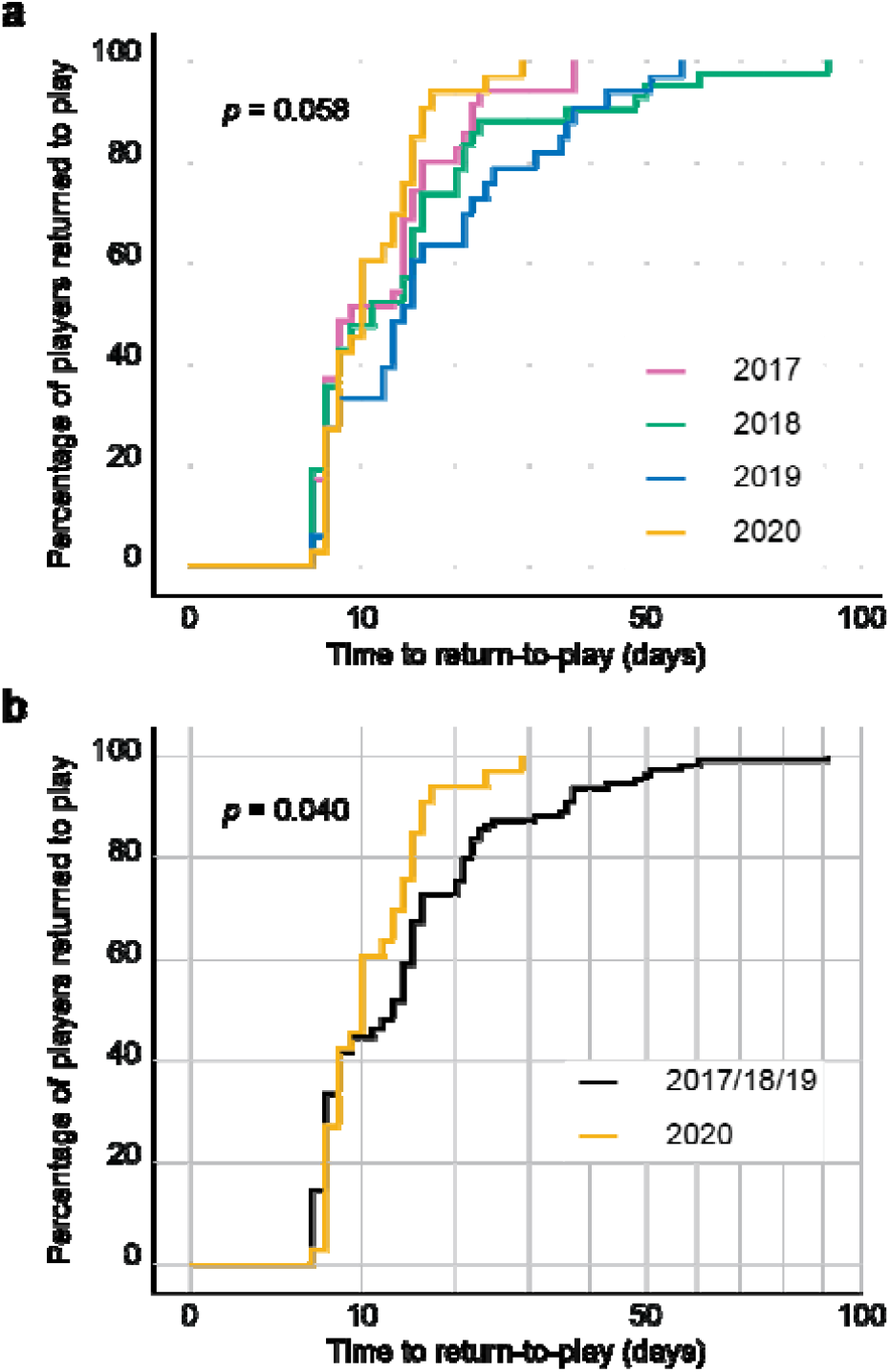
Time until return-to-play curves for (**a**) each year and (**b**) 2020 vs 2017-2019 combined. Mantel–Cox log rank test reveals 2020 has a significantly faster trajectory to return-to-play than other years combined (*p* = 0.040). (2017 *n* = 35, 2018 *n* = 42, 2019 *n* = 33, 2020 *n* = 33).

Taken together, these analyses of AFL public data demonstrate that the change in concussion management policy in 2020 has had no effect on the time to return-to-play. Paradoxically, return-to-play times in 2020 were found to be shorter than in the previous three years.

## 4.0 Discussion

In this study we examined public data to evaluate the effectiveness of an AFL policy change around concussion management. Under the new policy, the minimum interval from concussion to return-to-play would be expected to increase from seven to 12 days (to account for the five day extra medical clearance timespan) but the data shows that this has not occurred. In 2020, the mean number of rounds and days missed after concussion decreased, as opposed to increasing, and the overall trajectory of return-to-play was faster in 2020 than in the three years prior combined. Further we found that a high proportion of concussed players returned to competition in 2020 without missing a single game (44%), or within 10 days (56%) of a concussion.

These 2020 data stand in contrast to the ongoing improvements in the conservative management of concussion in the AFL from 2017 to 2019, as is clear from our data analyses as well as from the official injury report [11]. While the rationale for making the policy change is understood, its lack of effectiveness is more difficult to understand or explain. It may be due, at least in part, to the effects of COVID-19 restrictions and the associated stress in 2020, or it may be due to other factors affecting all years such as concomitant injuries, or the severity of concussion. However in the latter case, concussion is no longer graded by severity [12], so if a player is placed on the injury list with concussion, the minimum respite from play applies irrespective of the severity of injury. In some recent studies the mean symptom free duration from concussion injury was 28 days yet these players returned to sport in less than half that time [13, 14].

It is well described that medical staff can be pressured to clear players to return to competition as quickly as possible, and this pressure comes from multiple sources including the athlete themselves [15, 16]. In the extraordinary year of 2020, these pressures may have been amplified, thus leading to a shorter than average return-to-play time. But this does not excuse the possibility that many players with a concussion in 2020 have been prematurely returned to the game, risking further injury and the potential post-concussion effects. The reliance on athlete self-reporting on symptom resolution, observation from the doctor are clearly a ‘weak link’ in the staged return-to-play protocol, and this highlights the dire need for standardised objective measures of recovery after concussion. In the absence of such measures, sporting codes should be erring on the side of caution and mandating that at least one match be missed after suffering a concussion.

The reliance of this study on publicly available data provided by the AFL clubs themselves is a potential limitation. While we are not able to independently verify each case, the data are likely to be accurate as AFL clubs are thought to operate transparently in the listing of concussed players in the spirit of the sport, and they are unique among football codes in Australia in this regard. Also, state league data is not readily available to the public to assess whether players were returning to play *via* this pathway, which could have reduced the number of matches and days missed in previous years (due to COVID-19 this pathway did not operate in 2020). Despite this, we would still have expected to see increased time out of sport in 2020 due to the reduced opportunities for players to return to the AFL via other leagues. Conversely, if players in 2017-19 were returning within the week to compete in state leagues in spite of the consensus guidelines, this is still of concern with regards to return-to-play policies.

While return-to-play decisions are a shared decision-making process between the doctor, coach, and athlete across injuries generally [17], with concussion specifically the process must take into account whether an athlete’s decision-making capacity is compromised [17]. This should lay solely within the judgement of the clubs’ doctor, and ideally approached by objective means.

In conclusion our study demonstrates that the new AFL policy to increase rest and recovery time after concussion has not been effective. While there is good intention in addressing the ongoing concerns regarding concussion injuries in contact sports, improved monitoring methods during recovery and rehabilitation will contribute to compliance to the policy, and consequently long-term player health.

## Data Availability

All data is available from public websites as outlined in this manuscript.

## Acknowledgements

This research did not receive any specific grant from funding agencies in the public, commercial, or not-for-profit sectors. AJP currently receives partial research salary funding from Sports Health Check charity (Australia) and Erasmus+ strategic partnerships program (2019-1-IE01-KA202-051555). AJP has previously received partial research funding from the Australian Football League, Impact Technologies Inc., and Samsung Corporation, and has provided expert reports in concussion legal proceedings. No other author has any declaration of interest.

## Ethics approval

Waiver from Health and Disability Ethics Committee NZ

## Consent to participate

Not applicable

## Consent for publication

Not applicable

## Availability of Data and Material

All data is available from public websites as outlined in this manuscript.

## Code availability

Not applicable

## Authors’ contributions

AJP and CMS conceptualised the study, collected and analysed the data and contributed to the writing of the manuscript. DAK and AJW contributed to the writing and editing of the manuscript.

